# High-Density Lipoprotein (HDL) Subtypes Adversely Alter Brain Structure in Mild Cognitive Impairment: A Tensor-Based Morphometry Analysis

**DOI:** 10.1101/2024.08.20.24312114

**Authors:** Ali Azargoonjahromi, Mitra Ashrafi, Donya Abroushan, Elham Ramezannezhad, Mohammad Sadeghi, Seyede Roxane Pooresmaeil Niaki, Mehrsa Radmanesh, Amin Haratian, Azin Taki, Negar Nekahi, Yeganeh Moshiri, Marzieh Rahimi, Heidar Fadavian, Seyede Maryam Mousavi, Atousa Moghadam Fard, Mahsa Mayeli

**Author notes:** **Corresponding Authors:** Ali Azargoonjahromi, Shiraz University of Medical Sciences, Shiraz, Iran.

## Abstract

High-density lipoprotein (HDL) cholesterol is typically protective for cognitive function due to its anti-inflammatory, antioxidant, and vascular health benefits. However, recent studies indicated that certain HDL subtypes might be associated with adverse brain structural changes, commonly seen in mild cognitive impairment (MCI). Thus, further research is needed to understand the intricate relationship between HDL levels and brain structure, potentially leading to more effective therapeutic strategies. The current study aimed to investigate the impact of HDL subtypes, such as XL_HDL_P, M_HDL_FC_PCT, M_HDL_P, M_HDL_C, and M_HDL_CE, as well as APOA1, on brain structure in individuals with MCI using tensor-based morphometry (TBM). The study analyzed ADNI data from subjects with at least two serial MRI scans, processed using the Mayo TBM-Symmetric Normalization (SyN) pipeline and SyN for longitudinal measures. The CDR and ADAS scores were used to assess the severity of cognitive impairment and disease progression in our study participants. Significant ROIs were identified from a Mayo Clinic training set, and TBM-SyN scores were computed. The significant correlation was considered with p-values less than 0.05. The study found significant negative effects of several lipoproteins on TBM scores in individuals with MCI. Specifically, XL_HDL_P, with an effect size of - 0.00145 (p=0.029), and M_HDL_FC_PCT, with an effect size of -0.00199 (p=0.0016), were linked to lower TBM scores. Similarly, M_HDL_P (−0.00138, p=0.028), M_HDL_C (−0.00140, p=0.025), M_HDL_CE (−0.00136, p=0.031), and APOA1 (−0.00149, p=0.017) also showed significant associations. These findings indicate that higher levels of HDL subtype cholesterol are significantly associated with reduced TBM scores, suggesting that elevated levels are linked to adverse structural brain changes, such as atrophy, in individuals with MCI, potentially contributing to cognitive decline.

## 1. Introduction

High-density lipoprotein (HDL) is a type of lipoprotein that helps transport cholesterol from the bloodstream and tissues back to the liver for excretion or reuse (1). Often referred to as “good” cholesterol, HDL plays a key role in cardiovascular health by reducing the risk of atherosclerosis and other heart-related diseases (2, 3). Structurally, HDL particles are composed of a core of cholesterol and triglycerides encased in a layer of phospholipids and proteins, primarily apolipoprotein A-I (ApoA-I). This structure enables HDL to efficiently transport cholesterol from peripheral tissues and arterial walls back to the liver, a process known as reverse cholesterol transport (4, 5). Notably, HDL subtypes such as XL_HDL_P, M_HDL_P, M_HDL_C, and M_HDL_CE, indicate different sizes and concentrations of HDL particles, with larger ones being more effective in cholesterol transport (6, 7).

HDL is well-known for its cardiovascular benefits (8-10), but its importance in brain health is increasingly recognized. In the brain, cholesterol is not merely a structural component; it is essential for the formation and maintenance of neuronal membranes, synapses, and myelin sheaths (11-13). Indeed, HDL plays a crucial role in cholesterol homeostasis within the brain, facilitating its transport across the blood-brain barrier (BBB) to ensure neurons receive the precise amount needed for optimal function (14-17). Accordingly, by carrying newly synthesized cholesterol from astrocytes to neurons, HDL supports essential lipid-related physiological functions, synaptic maintenance, and plasticity, which are vital for learning and memory (14, 18).

In addition to its role in cholesterol transport, HDL exhibited significant anti-inflammatory (19-21) and antioxidant properties (22, 23) that are instrumental in mitigating neuroinflammation and oxidative stress—factors commonly associated with neurocognitive diseases such as mild cognitive impairment (MCI) and Alzheimer’s disease (AD) (24, 25). This means that HDL contributes to neuronal protection and disease progression attenuation by decreasing pro-inflammatory cytokines, reactive oxygen species, and potentially reducing amyloid-beta (Aβ) plaque accumulation. Elevated HDL levels are thus correlated with improved cognitive performance and a diminished risk of cognitive decline (19-23).

Nonetheless, recent research indicated that HDL’s impact on cognitive function is more nuanced than previously understood. For instance, dysfunctional HDL, resulting from oxidative modifications, can impair neuronal protection and potentially contribute to neurodegenerative conditions (26). Moreover, excessively high HDL levels have been associated with cognitive decline in certain studies, possibly due to intricate interactions with other lipoproteins and inflammatory pathways (27-29). In the context of AD, HDL’s role is complex: While it aids in the clearance of Aβ plaques (30, 31), some HDL subtypes may inadvertently exacerbate disease pathology (32, 33). Consequently, HDL’s effects on brain functions are contingent upon its functional state and the broader pathological environment.

Tensor-based morphometry (TBM) is a neuroimaging technique that analyzes brain structure by detecting subtle changes in tissue deformation from MRI scans. TBM is particularly effective in longitudinal studies, enabling the tracking of structural changes in the brain over time (34). This capability is crucial for elucidating HDL’s impact on brain health across different stages of aging or disease progression. Further, TBM provides regional specificity (35), allowing researchers to precisely identify how HDL affects specific brain regions associated with cognitive functions.

Thus far, and to the best of our knowledge, no research has specifically utilized TBM to investigate the impact of HDL on brain structure. Indeed, even though TBM has been extensively utilized to study brain atrophy in neurodegenerative diseases (34, 36-38), it has not yet been used to explore the role of HDL in this context; not to mention that structural atrophy in the brain often correlates with cognitive decline, yet it is not a definitive measure of cognitive impairment on its own.

Former research in related areas has utilized neuroimaging techniques like MRI to explore the relationship between cholesterol levels—including HDL—and brain structure. These studies, albeit with conflicting findings, have generally focused on how cholesterol affects brain volume, white matter integrity, and the progression of neurodegenerative diseases like AD. For example, some studies found correlations between lower HDL levels and reduced gray matter volume in brain regions crucial for cognitive function (39-41). This suggests that HDL might play a role in maintaining brain structure, though TBM, which can provide even more detailed structural analysis, has not yet been specifically employed for this purpose. Meanwhile, research using MRI-DTI pointed out that higher HDL levels are associated with better white and grey matter integrity, indicating that HDL can ascribe to preserving the structural health of the brain’s communication pathways (42-45).

The current study primarily aimed to address conflicting findings regarding HDL’s impact on brain structure among patients with MCI by employing TBM—a method that has not yet been utilized for this purpose. This study indeed aimed to investigate longitudinal structural changes in the brain and elucidate how HDL affects specific brain regions associated with neurocognitive disorders.

## 2. Methods and Materials

The data for this study were obtained from the Alzheimer’s Disease Neuroimaging Initiative (ADNI) database (http://adni.loni.usc.edu), a collaborative project established in 2003 under Dr. Michael W. Weiner. ADNI’s goal is to assess whether MRI, PET scans, biological markers, and clinical/neuropsychological evaluations can effectively track the progression of MCI and early AD. Participants, aged 55-90, underwent neuroimaging, lumbar punctures, and regular follow-ups, with detailed inclusion and exclusion criteria outlined elsewhere. Notable exclusions included a Hachinski Ischemic Score above 4, use of non-approved medications, recent changes in allowed medications, a Geriatric Depression Scale score of 6 or higher, and less than six years of education or equivalent work experience. In this study, 93 participants were classified into the MCI group according to ADNI’s clinical criteria.

### 2.1. Cognitive Assessment

The cognitive status of participants was evaluated using standardized assessments like the Alzheimer’s Disease Assessment Scale-Cognitive Subscale (ADAS-Cog) and the Clinical Dementia Rating (CDR) scale, which are essential tools for monitoring the progression of AD and related conditions.

The ADAS-Cog (46, 47) was specifically designed to measure the severity of cognitive symptoms associated with AD. It includes a series of cognitive tests that assess various domains such as memory, language, and praxis (motor coordination). Participants were asked to perform tasks like word recall, object naming, and copying geometric figures. The total score on the ADAS-Cog can range from 0 to 70, with higher scores indicating more severe cognitive impairment. This longitudinal tracking was crucial for understanding how cognitive decline progresses in individuals with MCI or AD (47).

The CDR (48) scale was used to assess the overall severity of dementia symptoms across multiple domains, including memory, orientation, judgment, and personal care. The evaluation process involved a semi-structured interview with the participant and an interview with an informant, typically a family member or caregiver. Based on these interviews, a clinician rated each domain on a scale from 0 (no impairment) to 3 (severe impairment). The ratings from each domain were then combined to produce a global CDR score, which classifies participants into categories such as no dementia, very mild dementia, mild dementia, moderate dementia, or severe dementia. The CDR score helps in classifying participants according to the stage of cognitive decline and is used to monitor changes in dementia severity over time (49).

### 2.2. Neuroimaging Processes

Longitudinal MRI measurements, crucial for detecting neurodegenerative changes in AD and used in clinical trials, led to the development of a robust, bias-free MRI metric for longitudinal studies. The Symmetric Diffeomorphic Image Normalization method was applied to normalize serial scans, producing TBM maps. Summary TBM-Symmetric Normalization (SyN) scores were calculated for each subject at follow-up time points by determining the SyN deformations between each follow-up and baseline scan. The software tools used included MATLAB, ANTs 1.9.x, and SPM5.

#### 2.2.1. Image Preprocessing for Each Individual Image

For each subject, we began with the “N3m” preprocessed datasets and created brain and ventricle masks for each image set. We generated an initial mean image from all N3m images and used SPM5-based mutual information co-registration, iteratively registering each N3m image to the mean. This process continued until the mean image stabilized or reached a maximum of 10 iterations. After the final iteration, all images were co-registered to the base image to ensure accuracy. We then used dilation, hole filling, and subtraction on the co-registered masks to isolate voxels dominated by white matter and cerebrospinal fluid (CSF), fitting Gaussian functions to the intensity spectra and scaling image intensities to standard values.

Next, using Aladin, we rigidly co-registered each image to the subject’s baseline image, averaged transformations within the subject, and resampled images and masks into this average space at 1mm isotropic resolution. We formed a new registration target by averaging the resampled images and masks. Affine registration between unregistered images and the average image was performed, followed by resampling into the target space. Finally, we balanced intensities and performed differential bias correction (DBC). Gaussian fits were used to determine white matter and CSF peak intensities for both the mean image and resampled images, with a GM-enhanced intensity spectrum calculated for gray matter. The gray matter, white matter, and CSF intensities were aligned with the mean image using spline-based intensity remapping. DBC was performed using voxels near CSF and white matter peak intensities, creating a dense field via tri-linear interpolation, smoothing it with a 20mm Gaussian kernel, and applying the result to achieve the final preprocessed image.

#### 2.2.2. Longitudinal Measure Free of Bias

High accuracy is often prioritized in warping algorithms, but asymmetric registration between serial scans can introduce bias in longitudinal measurements. We used the SyN algorithm, known for its symmetric registration and high accuracy, to compute deformations between preprocessed scans for each subject. We generated “annualized” log Jacobian maps by dividing log Jacobian voxels by the intrascan time interval. These deformations were applied to create “soft-mean” images, which were then segmented using SPM5, with ROI masks propagated to obtain mean annualized log Jacobian measurements in various ROIs.

#### 2.2.3. Mayo Clinic Patients for Region Selection

In developing longitudinal measurements, statistically significant ROIs are typically identified by analyzing a training set of patients and matched controls to capture neurodegenerative changes. In this study, we selected a training set of 51 AD subjects and 51 PiB-negative CN subjects matched by age, gender, and education, all with longitudinal MRI scans from the Mayo Clinic. To maintain a clean dataset, subjects had to maintain the same clinical diagnosis across both scans, with a baseline age of ≥ 64 years. Using a two-sample t-test, we selected the top 20 regions showing significant differences in GM volumes and longitudinal annualized log Jacobian data, resulting in 31 unique ROIs. Since 30 were GM ROIs (indicating shrinkage) and one was the ventricle (indicating expansion), we inverted the ventricle’s log Jacobian determinant before combining it with the GM values.

### 2.3. Lipoproteins Measurement

In this study, lipoprotein subtypes like HDL were quantified using Nightingale Health’s NMR metabolomics platform, which measured over 220 metabolic biomarkers from a single blood sample. This platform provided absolute concentrations of lipids and metabolites, enhancing data interpretability compared to relative measurements from mass spectrometry. Serum samples were prepared and measured between May 31st and June 9th. After thawing, the samples were mixed, centrifuged, and prepared using an automated liquid handler with a specific buffer solution. Measurements were taken with a Bruker AVANCE III HD 500 MHz spectrometer equipped with advanced NMR technology. The NMR data were processed and analyzed using Nightingale’s proprietary software, ensuring that metabolite levels were consistent with general population distributions and indicating high sample quality.

### 2.4. Statistical Analysis

The study utilized R and Python version 3.11 for statistical analysis. Feature selection was performed using F-regression analysis to identify significant metabolites related to brain structural changes. Metabolites with high F-statistics and significant p-values were selected for further analysis. A linear mixed-effects model, implemented in both R and Python, examined the relationship between selected metabolites and TBM imaging data, accounting for random effects at the individual level. Additionally, the study evaluated the association between metabolites and cognitive test scores. Effect sizes were calculated by comparing the R^2^ values of each metabolite’s model to a baseline model, revealing their contributions to structural brain alterations observed in TBM findings.

## 3. Results

The study recruited 93 patients diagnosed with MCI. The cohort had a mean age of 65.23 years (±6.47). Cognitive function was assessed using the ADAS-Cog and CDR scales, with mean scores of 8.31 (±4.97) and 0.42 (±0.30), respectively. Among the participants, 54.8% (n=51) were male and 45.2% (n=42) were female (Table 1).

**Table 1.:** Demographic characteristics of the participants.

| Number of MCI Subjects |  | Mean age of the patients | Mean ADAS-Cog score | Mean CDR score |
| --- | --- | --- | --- | --- |
| N= 93 | | 65.23 years ($\pm 6.47$) | 8.31 ($\pm 4.97$) | 0.42 ($\pm 0.30$) |
| Male | Female |  |  |  |
| 54.8% (n= 51) | 45.2% (n= 42) |  |  |  |

The TBM analysis revealed several significant relationships between HDL subtypes and brain structural changes. For instance, XL_HDL_P exhibited a fixed effect estimate of -0.00145 with a p-value of 0.0293, indicating a significant impact on brain structure with a modest effect size (R^2^ = 0.0132). This suggests that higher levels of XL_HDL_P are associated with reduced brain tissue volume or alterations in brain density. Similarly, M_HDL_FC_PCT showed a strong fixed effect estimate of -0.00199 and a highly significant p-value of 0.0016, reflecting a substantial association with brain structural changes, likely indicating pronounced atrophy or changes in brain shape, as evidenced by a larger effect size of 0.0292.

M_HDL_P also demonstrated a significant relationship, with a fixed effect estimate of -0.00138 and a p-value of 0.0276, which implies a moderate impact on brain structure, potentially related to localized atrophy or changes in brain density. Likewise, M_HDL_C had a fixed effect estimate of -0.00140 and a p-value of 0.0254, showing a significant but slightly smaller effect size of 0.0155. M_HDL_CE presented a fixed effect estimate of -0.00136 with a p-value of 0.0306, indicating a significant association with modest structural changes or density reductions, with an effect size of 0.0145. Additionally, APOA1 had a fixed effect estimate of -0.00149 and a p-value of 0.0171, reflecting a notable impact on brain structure, with an effect size of 0.0181 (Table 2).

**Table 2.:**
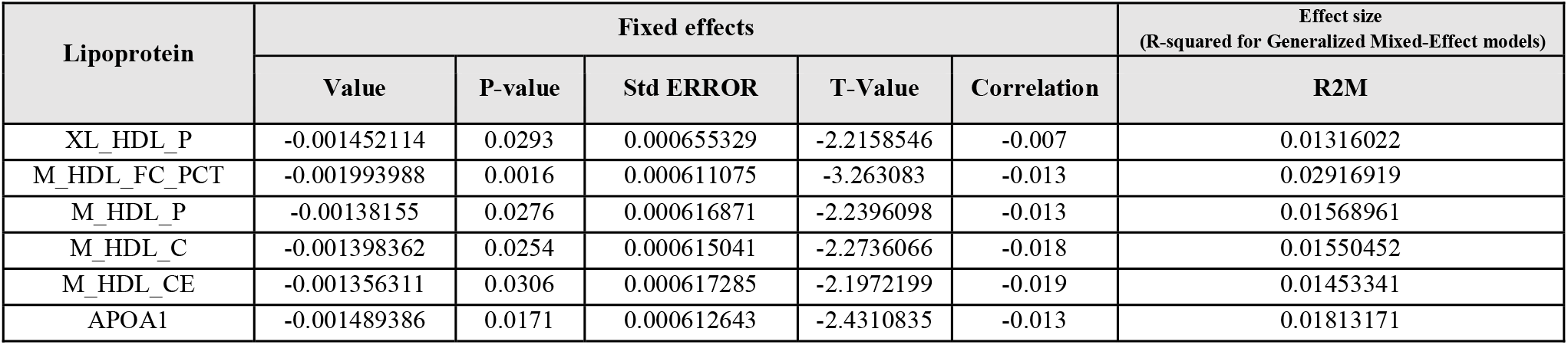
Statistical analysis of HDL subtypes associations with brain structural changes using TBM.

| Lipoprotein | Fixed effects |  |  |  |  | Effect size<br>(R-squared for Generalized Mixed-Effect models) |
| --- | --- | --- | --- | --- | --- | --- |
|  | Value | P-value | Std ERROR | T-Value | Correlation | R2M |
| XL_HDL_P | -0.001452114 | 0.0293 | 0.000655329 | -2.2158546 | -0.007 | 0.01316022 |
| M_HDL_FC_PCT | -0.001993988 | 0.0016 | 0.000611075 | -3.263083 | -0.013 | 0.02916919 |
| M_HDL_P | -0.00138155 | 0.0276 | 0.000616871 | -2.2396098 | -0.013 | 0.01568961 |
| M_HDL_C | -0.001398362 | 0.0254 | 0.000615041 | -2.2736066 | -0.018 | 0.01550452 |
| M_HDL_CE | -0.001356311 | 0.0306 | 0.000617285 | -2.1972199 | -0.019 | 0.01453341 |
| APOA1 | -0.001489386 | 0.0171 | 0.000612643 | -2.4310835 | -0.013 | 0.01813171 |

Overall, all the lipoproteins analyzed in the TBM study—XL_HDL_P, M_HDL_FC_PCT, M_HDL_P, M_HDL_C, M_HDL_CE, and APOA1—were found to be statistically significant, with p-values below the standard threshold of 0.05. This indicates that each of these lipoproteins has a notable association with brain structural changes, including reductions in brain volume (atrophy), decreases in tissue density, and alterations in brain shape, reflecting their potential impact on brain health.

## 4. Discussion

The current study, based on our knowledge, is the first to report the use of TBM to investigate the role of HDL subtypes and APOA1 in brain structure and formation. We found that higher levels of XL_HDL_P, M_HDL_FC_PCT, M_HDL_P, M_HDL_C, and M_HDL_CE, as well as APOA1 are linked to reductions in brain volume, decreases in tissue density, and alterations in brain shape. These findings suggested that elevated levels of these lipoproteins may contribute to brain atrophy and other structural changes that are often associated with cognitive decline and neurodegenerative conditions.

Consistent with our findings, few studies have also reported that elevated HDL levels may be associated with an increased risk of neurocognitive disorders. For example, a study analyzing data from the ASPREE trial found that older adults with plasma HDL-C levels above 80 mg/dL had a 27% higher risk of developing dementia over an average follow-up of 6.3 years, especially in those aged 75 and older. This association remained significant even after adjusting for factors such as age, sex, and APOE genotype, suggesting that high HDL-C levels could be a risk factor for dementia in older adults (32). Similarly, a study from the China Health and Retirement Longitudinal Study (CHARLS) revealed that higher variability in HDL-C levels was linked to an increased risk of cognitive decline. In a cohort of 5,930 participants, those with the highest HDL-C variability had a greater likelihood of cognitive decline compared to those with the lowest variability. The findings suggest that reducing HDL-C variability might help lower the risk of cognitive decline in the general population (28).

In addition, recent studies suggest that very high HDL-C levels may increase the risk of dementia, even in younger individuals with comorbidities (50). Elevated HDL has also been linked to poorer cognitive recovery after a stroke, possibly due to metabolic stress (51). In Parkinson’s disease, higher HDL levels are associated with cognitive decline, particularly in women. These findings indicate that while HDL is generally considered heart-healthy, its impact on brain health may be more complex and potentially harmful in certain contexts (27).

The link between high HDL-C and increased dementia risk is not fully understood. While moderately high HDL-C levels have been associated with better cognitive function, very high HDL-C might be dysfunctional and not reflect effective lipid transport (52). It is also possible that both high HDL-C and dementia could stem from an unrelated pathology. With dementia becoming a growing concern in aging populations, easily measurable markers like HDL-C could be useful, though they may only apply to a small subset of individuals. Hence, further research could clarify the role of high HDL-C in dementia, potentially leading to new insights into its pathogenesis and related health risks.

Nonetheless, our findings contrast with those of a recently published meta-analysis of data from 100 studies suggesting that HDL-C levels were unrelated to dementia (53). Likewise, several studies collectively highlighted the significant role of HDL-C in cognitive function across various populations, including aging women (54), nonagenarians (55), centenarians (56), and patients with bipolar disorder (57), diabetes mellitus (58), and Parkinson’s disease (59). Higher HDL-C levels were consistently associated with better cognitive performance, particularly in areas like memory and learning in such studies. In some cases, low-normal total cholesterol (TC) levels were linked to reduced cognitive function and brain atrophy, suggesting a complex relationship between lipid levels and brain health (60). These findings underscore the protective effects of HDL-C on cognitive function and support further research into its potential as a therapeutic target for neurodegenerative diseases.

While the link between high HDL and increased dementia risk remains unclear, several mechanisms may be involved. Dysfunctional HDL at elevated levels could disrupt lipid transport, compromising neuronal membrane integrity and BBB function, which may lead to neuroinflammation and oxidative stress. Additionally, abnormal HDL could impair the clearance of Aβ peptides, promoting amyloid plaque accumulation in the brain. In short, these processes might contribute to structural brain changes associated with cognitive decline and dementia (26-29, 32, 33).

One of the main strengths of this study was its detailed analysis of HDL subtypes, which offered a more nuanced understanding of the relationship between specific lipoproteins and brain structure than studies that only considered overall HDL-C levels. This focus on subtypes allowed the research to explore the intricate ways in which different forms of HDL might influence brain health. Another significant strength was the use of advanced imaging techniques, specifically TBM with the Mayo TBM-SyN pipeline. This approach enabled precise measurement of structural brain changes, which enhanced the reliability and accuracy of the findings. Additionally, the study utilized longitudinal data, with serial MRI scans over time, which strengthened its ability to observe changes in brain structure in relation to HDL levels, providing a more dynamic view of how these factors interplayed over time. The relevance of the study to cognitive decline, particularly in individuals with MCI, was also notable, as it directly addressed a critical area in the understanding of AD progression.

However, the study also had some limitations. As an observational study, it could not establish a causal relationship between HDL subtypes and brain atrophy, meaning that the observed associations might have been influenced by other confounding factors. Another limitation was the potential lack of generalizability. Moreover, the study primarily focused on structural brain changes without directly assessing how these changes translated into cognitive or functional outcomes. This left an incomplete picture of the clinical relevance of the findings. Finally, although advanced imaging techniques were used, there was the potential for measurement variability. Differences in MRI scanners, protocols, or image processing methods could have introduced inconsistencies in the TBM scores across participants, potentially affecting the study’s results.

## 5. Conclusion

The study aimed to examine the impact of various HDL subtypes on brain structure in individuals with MCI using TBM. We found that higher levels of specific HDL subtypes, such as XL_HDL_P, M_HDL_FC_PCT, M_HDL_P, M_HDL_C, and M_HDL_CE, as well as APOA1, are significantly associated with lower TBM scores, indicating adverse structural changes like brain atrophy. These findings suggest that elevated levels of these HDL subtypes may contribute to cognitive decline by negatively affecting brain structure in MCI patients.

## Data Availability

The data used in this research was obtained from Alzheimer's Disease Neuroimaging Initiative (ADNI) and is available with permission to all researchers.

## Abbreviations

HDL: High-density lipoprotein
BBB: blood-brain barrier
MCI: mild cognitive impairment
AD: Alzheimer’s disease
Aβ: amyloid-beta
TBM: Tensor-based morphometry
ADAS-Cog: Alzheimer’s Disease Assessment Scale-Cognitive Subscale
CDR: Clinical Dementia Rating
CSF: cerebrospinal fluid
DBC: differential bias correction

## Declaration sections

## Acknowledgments

Data used in preparation of this article were obtained from the Alzheimer’s Disease Neuroimaging Initiative (ADNI) database (https://adni.loni.usc.edu/). As such, the investigators within the ADNI contributed to the design and implementation of ADNI and/or provided data but did not participate in analysis or writing of this report. A complete listing of ADNI investigators can be found at: http://adni.loni.usc.edu/wp-content/uploads/how_to_apply/ADNI_Acknowledgement_List.pdf.

## Ethical Approval and Consent to Participate

This study was conducted using ADNI data. The ADNI study is ethically approved and operated in accordance with the Declaration of Helsinki, 1964.

## Consent for Publication

Not applicable.

## Authors’ Contributions

All authors listed have made a substantial, direct, and intellectual contribution to the work, and approved it for publication. A.A., M.A., D.A., A.M.F., and E.R. contributed to developing research ideas and analysis. M.S., M.R., M.M., and A.T. contributed to interpretation of data, writing the draft, and revising it. S.R.P.N., A.H., and H.F. reanalyzed data and contributed to editing draft. N.N., Y.M., M.R., and S.M.M. contributed to writing the draft, and revising it. All authors read and approved the final manuscript.

